# Lab-wide association scan of polygenic scores identifies biomarkers of complex disease

**DOI:** 10.1101/2020.01.24.20018713

**Authors:** Jessica K Dennis, Julia M Sealock, Peter Straub, Donald Hucks, Ky’Era Actkins, Annika Faucon, Slavina B Goleva, Maria Nirachou, Kritika Singh, Theodore Morley, Douglas M. Ruderfer, Jonathan D Mosley, Guanhua Chen, Lea K Davis

## Abstract

Clinical laboratory (lab) tests are used in clinical practice to diagnose, treat, and monitor disease conditions. Test results are typically stored in electronic health records (EHRs), and a growing number of EHRs are linked to patient DNA, offering unprecedented opportunities to query relationships between clinical lab tests and genetics. Clinical lab data, however, are of uneven quality, and previous studies have focused on a small number of lab traits. We present two methods, QualityLab and LabWAS, to clean and analyze EHR labs at scale in a Lab-Wide Association Scan. In a proof of concept analysis focused on blood lipids and coronary artery disease, we found that heritability estimates of QualityLab lipid values were comparable to previous reports; polygenic scores for lipids were strongly associated with the referent lipid in a LabWAS; and a LabWAS of a polygenic score for coronary artery disease recapitulated known heart disease biomarker profiles and identified novel associations. Our methods extend previous EHR-based analysis tools and increase the amount of EHR data usable for discovery.

## Introduction

The overarching goal of this study was to determine whether laboratory (lab) test results collected in a hospital and outpatient setting could be mined against polygenic scores (PGS) to identify known and novel biomarker associations for complex disease. Lab test results are essential to routine clinical care. These targeted biochemical measurements facilitate disease diagnosis and influence health care delivery. Clinical lab values are also monitored as mediators of disease risk, and are targeted by interventions to reduce disease incidence (e.g., cholesterol-lowering medication to reduce the risk of heart disease). Lab test results in the electronic health record (EHR) are a vast and growing resource for novel biomarker discovery, especially as EHRs are increasingly linked to patient DNA samples (e.g., the eMERGE consortium (https://emerge.mc.vanderbilt.edu)), the All of Us Program (https://allofus.nih.gov), and the Million Veteran’s Program (https://www.research.va.gov/mvp/)). Genetic studies of EHR-based labs could reveal novel biomarker-disease or biomarker-gene associations, which in turn could lead to better understanding of biological processes in disease, improved diagnostic algorithms and new therapeutic targets.

Despite their potential, however, EHR-based labs have been used in only a handful of prior genetic studies^1-5^, and none have systematically interrogated an extended collection of EHR-based lab values. Barriers to studying EHR-based labs include uneven data quality, and challenges inherent to analyzing and interpreting high-dimensional health care data. Data entry errors exist, resulting in implausible recorded values^6^, some labs have different units and reference ranges over time, and many individuals have multiple observations of different lab tests, each measured at varying times relative to diagnoses and treatment^7^. Moreover, previous studies demonstrate that while 99% of lab results are accurately transmitted from the testing laboratory to the EHR, only 70% of test results contain all required reporting elements, and only 91% of results are appropriately formatted^8^. Thus, while these data represent real clinical care and may accelerate translational research, there is little precedent for their analysis and interpretation in genetic studies.

To address these challenges, we present a high-throughput framework for genetic analysis of EHR-derived lab data. We have developed two methods: the QualityLab pipeline to clean, standardize, and visualize lab data; and the Lab-Wide Association Scan (LabWAS) pipeline to scan for associations between any variable of interest (genetic or otherwise) and the cleaned EHR labs. The LabWAS method is similar to the Phenome-Wide Association Scan (PheWAS) which scans for association between an exposure variable (typically, a genetic risk factor) and many phenotypes^9^. The PheWAS method has replicated many known gene-disease associations^10^ and has identified novel pleiotropic genetic effects^11^, opportunities for drug repurposing, and unintended drug consequences^12^.

We hypothesized that EHR-based lab values could be used to identify known and novel relationships between genetics, biomarkers, and disease. We deployed our framework in the Vanderbilt University Medical Center (VUMC) EHR and linked biobank, BioVU, and focused on genetic analysis of blood values of high-density lipoprotein cholesterol (HDL), low-density lipoprotein cholesterol (LDL), and triglycerides (TG), and on coronary artery disease (CAD) as proof-of-principle examples to test the association between PGS for CAD and known biomarkers of disease (LDL, HDL, and TG) using the QualityLab and LabWAS methods. We show that EHR-derived lipids values are genetically similar to those in population-based studies, and that PGS for lipids robustly associate with their respective lab in a LabWAS. Additionally, our LabWAS revealed that PGS for CAD associated with known lipid biomarkers and with potentially novel immune biomarkers.

## Methods

### Study Sample

VUMC is a tertiary care center that provides inpatient and outpatient care in Nashville, TN. The VUMC EHR was established in 1990 and includes data on billing codes from the International Classification of Diseases, 9th and 10^th^ editions (ICD-9 and ICD-10), Current Procedural Terminology (CPT) codes, laboratory values, reports, and clinical documentation. The de-identified mirror of the EHR, known as the Synthetic Derivative, includes patient records on more than 2.8 million individuals. In 2007, VUMC launched a biobank, BioVU, and the BioVU Consent form is provided to patients in the outpatient clinic environments at VUMC. The form states policies on data sharing and privacy, and upon consent, makes any blood leftover from clinical care eligible for BioVU banking^13^. The VUMC Institutional Review Board oversees BioVU and approved this project.

### Genotyping and Quality Control

We obtained genotype information on 94,474 BioVU individuals genotyped on the Illumina MEGA^EX^ array. Using PLINK v1.9^14^ genotypes were filtered for SNP and individual call rates, sex discrepancies, and excessive heterozygosity (Supplementary Material). We selected individuals of European ancestry using principal component analysis implemented in Eigenstrat^15,16^ and confirmed the absence of genotyping batch effects through logistic regression with ‘batch’ as the phenotype. Imputation was completed using the Michigan Imputation Server^17^ using the Haplotype Reference Consortium (HRC) reference panel. SNPs were then filtered for SNP imputation quality (R^2^>0.3) and converted to hard calls. We restricted to autosomal SNPs, filtered SNPs with minor allele frequency >0.01, or with allele frequencies that differed by more than 10% from the 1000 Genomes Project phase 3 CEU set^18^, and Hardy-Weinberg Equilibrium (p>1×10^−10^). The resulting dataset contained 9,386,383 SNPs on 72,828 individuals of European genetic ancestry.

### QualityLab Pipeline

In parallel with the BioVU genotyping project, we extracted data on all lab tests collected in the routine clinical care of 70,704 BioVU patients, amounting to 59,463,045 observations across 6,407 lab tests (Figure 1a). Of these lab tests, 2,865 were reported in non-numeric values and 467 had only been administered to one patient, leaving 3,075 quantitative lab tests for further cleaning. Some lab tests had observations recorded in different units (e.g., Selenium reported in both mcg/L and ug/L), thus we restricted to lab tests for which at least 70% of the observations were measured in the same unit and required that each lab have at least 100 patients and at least 1,000 numeric observations, for a total of 481 labs retained for further analysis.

**Figure 1.**
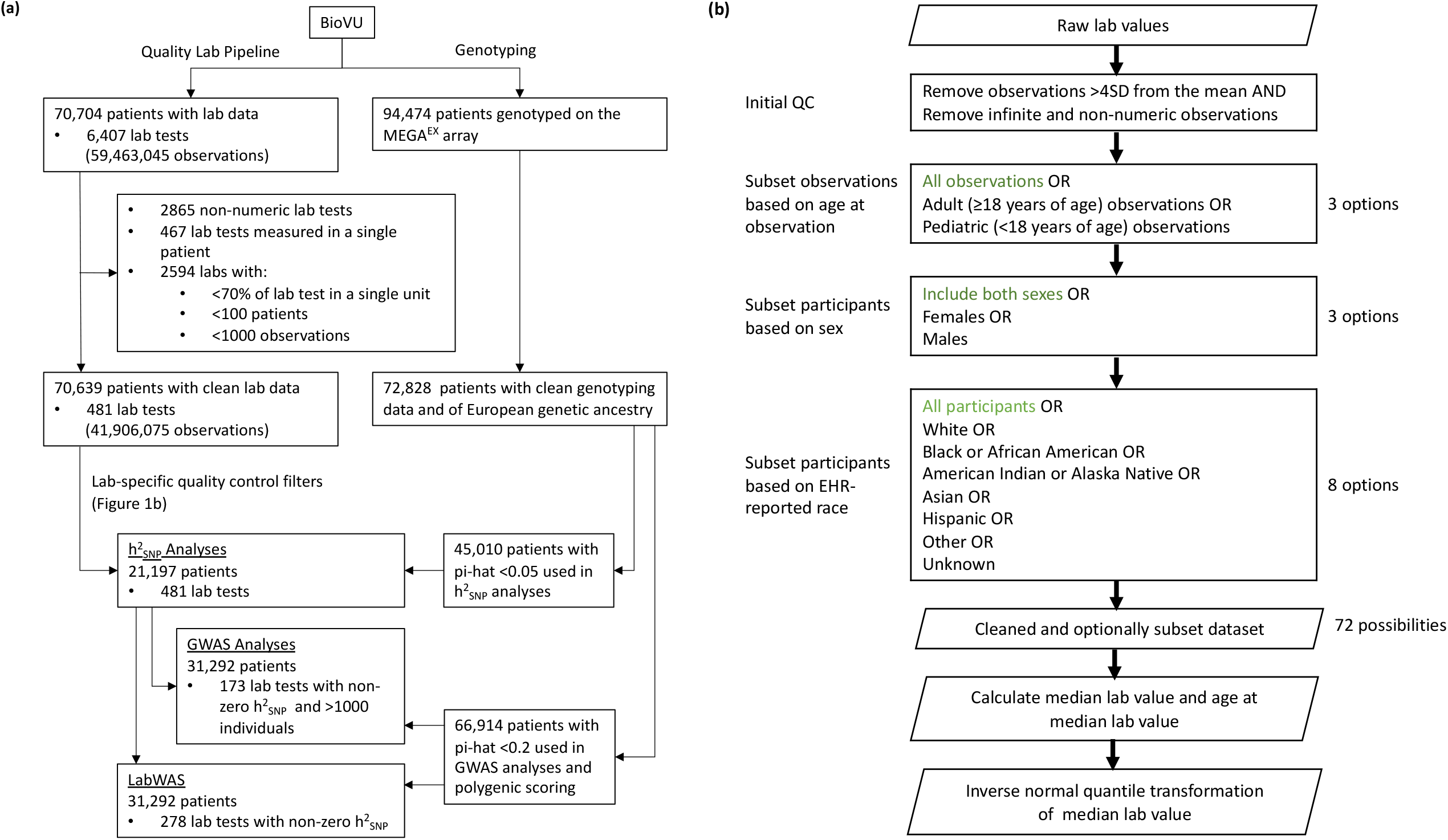
Selection of BioVU patients and datasets for different analyses presented in this manuscript. (a) BioVU patients were selected in parallel for clinical laboratory (lab) test cleaning, and for genotyping. (b) Lab-specific quality control filters and subsetting were applied to the 481 lab tests in the 70,639 patients with clean lab data. Parallelograms denote input and output datasets. Options highlighted in green were selected for the proof-of-principle analyses of blood-based lipid lab values.

For each of these 481 labs, we applied lab-specific quality control filters (Figure 1b). First, we filtered infinite and non-numeric values, as well as observations outside of 4 standard deviations from the overall sample mean, indicative of biologically implausible values due to technical or recording errors, monogenic disorders, or extreme environmental influence. We calculated the median lab value for each patient and extracted the patient’s age at median lab value. In patients with an even number of observations, we defined the age at median lab value as the mid-point of the patient’s ages at the two lab values on either side of the median lab value.

The analyses presented in this manuscript use the QualityLab dataset constructed from pediatric and adult observations, in both sexes, in patients of all races (Figure 1b). In downstream genetic analyses, however, we restrict to participants of European genetic ancestry to match the ancestry of the participants in the GWAS used for the construction of PGS.

The QualityLab pipeline also provides user with the option to stratify data (Figure 1b), by age at observation, sex, and EHR-recorded race, for a total of 72 different data subsets. The QualityLab pipeline generates summary statistics and plots for each strata (e.g., mean, maximum, and minimum of the median lab value; Supplementary Table 1; Supplementary Figure 1), and returns two versions of the data for downstream analyses. The first is a table of median lab values and age at median lab value for each individual. The second is an inverse normal quantile transformation (INT) of the median lab value data, to account for skewness and non-normality^19,20^. Importantly, the choice of quality control thresholds is completely in the control of the user. The choices made here reflect the goals of this study which focus on the central tendencies of large populations. However, the outlier thresholds and normalization methods employed here would not be appropriate in a study of rare, potentially pathogenic, variation where large genetic effects and extreme phenotypes may be expected.

### Heritability and GWAS Analyses

Prior to SNP-based heritability (h^2^_SNP_), we first calculated pairwise relatedness in the BioVU genotyped sample and randomly removed one related individual from pairs with pi-hat greater than 0.05, leaving 45,010 individuals of European genetic ancestry (Figure 1a). We then used the Genome-wide Complex Trait Analysis (GCTA) package (version 1.92.4)^21^ to create a genetic relationship matrix for all pairwise individuals, and heritabilities were calculated using restricted maximum likelihood (REML) methods. We used the median, INT-transformed lab values from the QualityLab pipeline, and of the 481 analyzed labs, 278 had non-zero heritability. For GWAS analyses, we randomly removed one related individual from pairs with pi-hat greater than 0.2 (n=66,914). Next, we subset to the heritable labs with at least 1,000 individuals (n=173), and ran a GWAS using fastGWA^22^ (Figure 1a). All h^2^_SNP_ and GWAS analyses included covariates for sex, cubic splines (knots=4) of median age across the medical record (to control for non-linear effects of age), and the top 10 principal components of ancestry.

### Heritability and GWAS Analyses of Lipids

We benchmarked our h^2^_SNP_ estimates against those from an external dataset, the Global Lipids Genetics Consortium (GLGC)^23^. GLGC estimates of h^2^_SNP_ for HDL, LDL, and TG were calculated from GWAS summary statistics using LDSC^24^. We computed h^2^_SNP_ in BioVU using Linkage Disequilibrium Score regression (LDSC) applied to our fastGWA summary statistics for HDL, LDL, and TG (Supplementary Figure 2). However, because LDSC can underestimate h^2^_SNP_25, we also calculated h^2^_SNP_ using GCTA. In addition to these h^2^_SNP_ comparisons, we calculated the genetic correlations (r_g_) between the BioVU lipid GWASs and the GLGC lipid GWAS using LDSC and the pre-computed European LD scores from 1000 Genomes Phase 3 European data^26^. In sensitivity analyses, we repeated genetic correlations of LDL after controlling the BioVU GWASs for coronary atherosclerosis or diabetes diagnoses, defined as phecodes 411, “Ischemic heart disease” and 249, “Secondary diabetes mellitus” (Supplementary Material).

To validate EHR-based lipid values, we tested the robustness of HDL, LDL, and TG h^2^_SNP_ estimates to different lab value and patient filters. First, we excluded lipid measurements that occurred after the first mention of lipid-altering mediation in the EHR (Supplementary Material), and re-calculated each patient’s pre-medication median values of HDL, LDL, and TG. Second, we excluded patients with a diagnosis of CAD, defined by the phecode 411 (Supplementary Material).

### LabWAS Pipeline

LabWAS uses the median, INT-transformed lab values from the QualityLab pipeline in a linear regression to determine the association with an input variable, adjusting for covariates. In these analyses, a primary goal of the LabWAS was to test common population genetic variation (e.g., PGS) for association with common population variation in lab values. We therefore only included the 278 labs with non-zero h^2^_SNP_.

### Polygenic Scoring

Prior to polygenic scoring, we randomly removed one related individual from pairs with pi-hat greater than 0.2, leaving 66,914 individuals of European genetic ancestry (Figure 1a). We generated PGS for these individuals using PRS-CS ^27^. PRS-CS is a recently developed Bayesian polygenic prediction method that imposes continuous shrinkage priors on SNP effect sizes (Polygenic Risk Score – Continuous Shrinkage)^27^. These priors can be represented as global-local scale mixtures of normals which allows the model to flexibly adapt to differing genetic architectures and provides substantial computational advantages. The shrinkage parameter was automatically learnt from the data (i.e., using PRS-CS-auto). SNP effect estimates were obtained from GWAS summary statistics and the score was calculated using a linkage disequilibrium reference panel from 503 European samples in the 1000 Genomes Project phase 3^18^. Although PRS-CS outperformed other polygenic scoring methods across a range of traits in previous experiments, its superiority may not hold across all genetic architectures ^27^. We therefore also generated polygenic scores using LDPred ^28^ and PRSice-2 ^29^ (Supplementary Material), and have automated a pipeline to generate scores across all three methods. PGS were scaled to have a mean of zero and SD of one before testing for association with any outcome variables. We validated each score by testing the proportion of trait variability explained by the PGS, controlling for sex, cubic splines of median age (4 knots) across the medical record, and the top 10 principal components to adjust for genetic ancestry (Supplementary Figure 5).

### LabWAS of Polygenic Scores

PGS for LDL (PGS_LDL_), HDL (PGS_HDL_), and TG (PGS_TG_), were calculated in BioVU participants using PRS-CS and applying SNP weights from the GLGC GWAS summary statistics. We then ran LabWAS of PGS_LDL_, PGS_HDL_, and PGS_TG_ to test whether lipid labs were robustly associated with the genetic scores to which they corresponded. Next, a PGS for CAD (CAD_PGS_) was calculated using SNP weights from CARDIoGRAMplusC4D GWAS summary statistics^30^ and a LabWAS of PGS_CAD_ to test whether the score could identify lab traits associated with genetic risk for CAD, before and after controlling for a CAD diagnosis (Supplementary Material). Each LabWAS was controlled for sex, cubic splines of median age across the medical record, and the top 10 principal components of ancestry. Results are reported as effect estimates and their 95% confidence intervals per SD increase in the polygenic score. The Bonferroni-corrected threshold for statistical significance across all tested labs was 1.80×10^−4^ (0.05/278).

## Results

### QualityLab Pipeline

The QualityLab pipeline identified 70,639 BioVU patients with clean lab data, of whom 31,292 were also of European genetic ancestry and were included in the polygenic score LabWAS analyses (Figure 1a). These 31,292 patients had data on 278 labs with non-zero h^2^_SNP_, containing 18,144,061 observations. The median number of unique lab tests per patient was 55, and the median number of lab observations per patient was 292. Slightly more than half of the BioVU patients in the sample were women (53.5%), and the average median age across the EHR was 53.7 years. These BioVU participants included 6,514 CAD cases and 15,886 CAD controls (Supplementary Material; Supplementary Table 3).

### Heritability and GWAS Analyses

Across all 481 clean lab traits, h^2^_SNP_ was non-zero in 278 labs and the p-value ranged from 2 × 10^−6^ to 0.94. (Supplementary Table 4, Supplementary Figure 2?). The GWAS summary statistics for the labs with calculable heritability and at least 1,000 individuals (n=173) are available here: https://www.dropbox.com/sh/w1pbe0jq1bjkpc5/AAAUIdtBgUybE6iHraE8jvp8a?dl=0.

### Heritability and GWAS Analyses of Lipids

The h^2^_SNP_ estimates in BioVU were robust to removing post-medication observations, and to removing CAD cases. The number of participants included in these analyses, however, was smaller, and so the standard errors of these h^2^_SNP_ estimates were larger (Figure 2a; Supplementary Table 5). Both GCTA and LDSC gave similar estimates of h^2^_SNP_ in BioVU (Figure 2b), and the LDSC estimates in BioVU were comparable to those in the GLGC for LDL, but less so for HDL and TG. Genetic correlation between BioVU and GLGC summary statistics was strong for HDL (rg=0.86, SE=0.12, p-value=5.42 × 10^−13^) and TG (rg=0.89, SE=0.06, p-value=2.16 × 10^−43^). However, genetic correlation for LDL was non-significant (rg=-0.01, SE=0.73, p-value=0.99). The genetic correlation increased when we restricted to pre-medication values of LDL in BioVU (rg=0.51, SE=0.48, p-value=0.29) (Figure 2c), and increased further when we controlled for coronary atherosclerosis and diabetes diagnoses (rg=0.81, SE=1.18, p-value=0.49). The power, however, remained low and the standard error remained large (Supplementary Figure 4).

**Figure 2.**
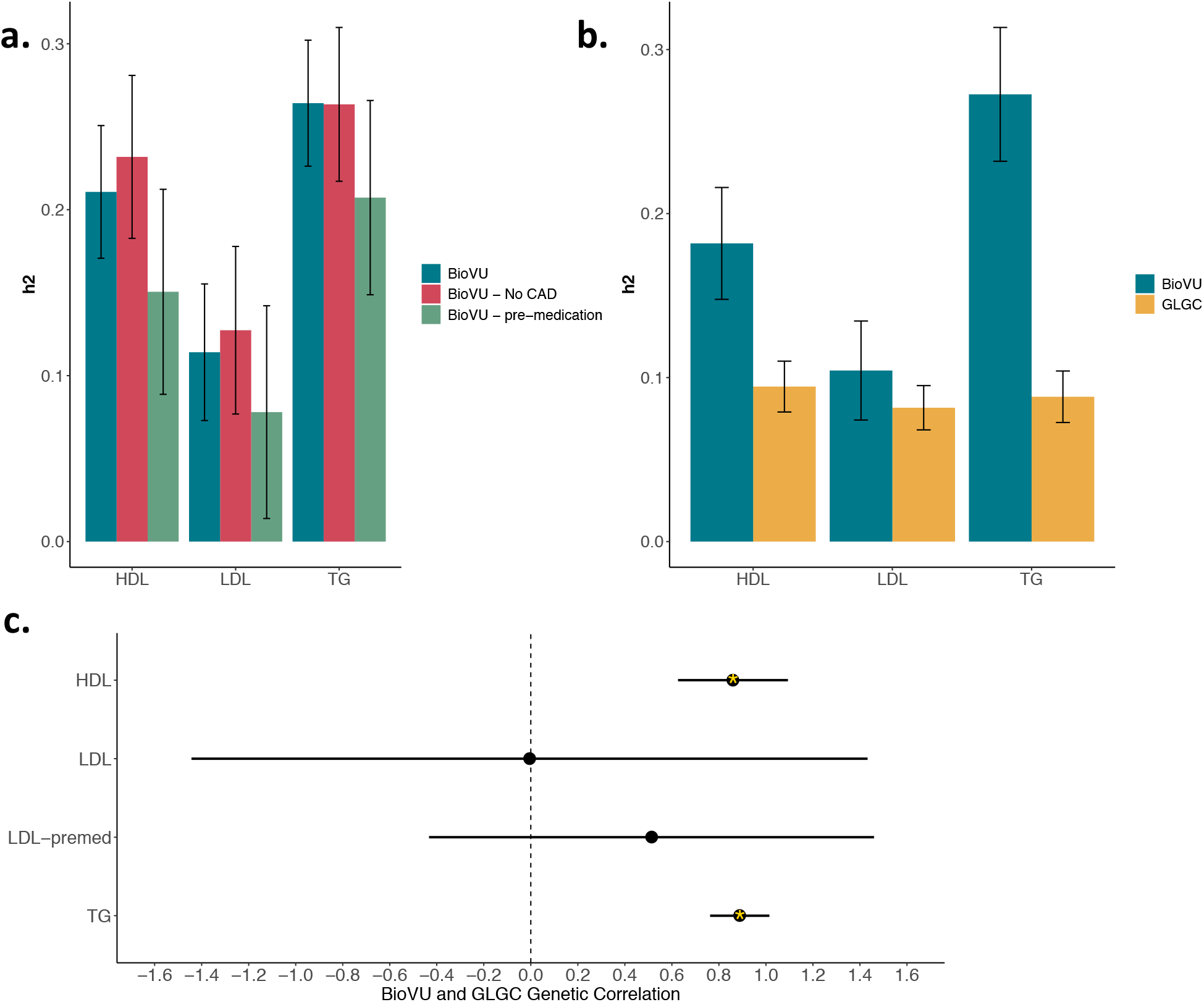
Heritability and GWAS analyses of lipids. (a) Estimates of heritability computed by GCTA in BioVU patients were robust to excluding individuals with a diagnosis of CAD, and to removing post-medication observations. (b) Estimates of heritability computed using GWAS summary statistics and LDSC were comparable across BioVU and the Global Lipids Genetic Consortium (GLGC) samples. (c) Genetic correlations between lipid levels in BioVU and the Global Lipids Genetic Consortium (GLGC). Stars denote statistically significant correlations.

### LabWAS of Polygenic Scores for Lipids

A LabWAS of HDLPGS was associated with levels of several metabolic markers (Figure 3a, Supplementary Table 6), including increased HDL, decreased TG, increased total blood cholesterol, and increased blood glucose. HDLPGS was also associated with five other lab values, including two blood composition labs (carbon dioxide and platelet count), two kidney related measurements, and one liver lab.

**Figure 3.**
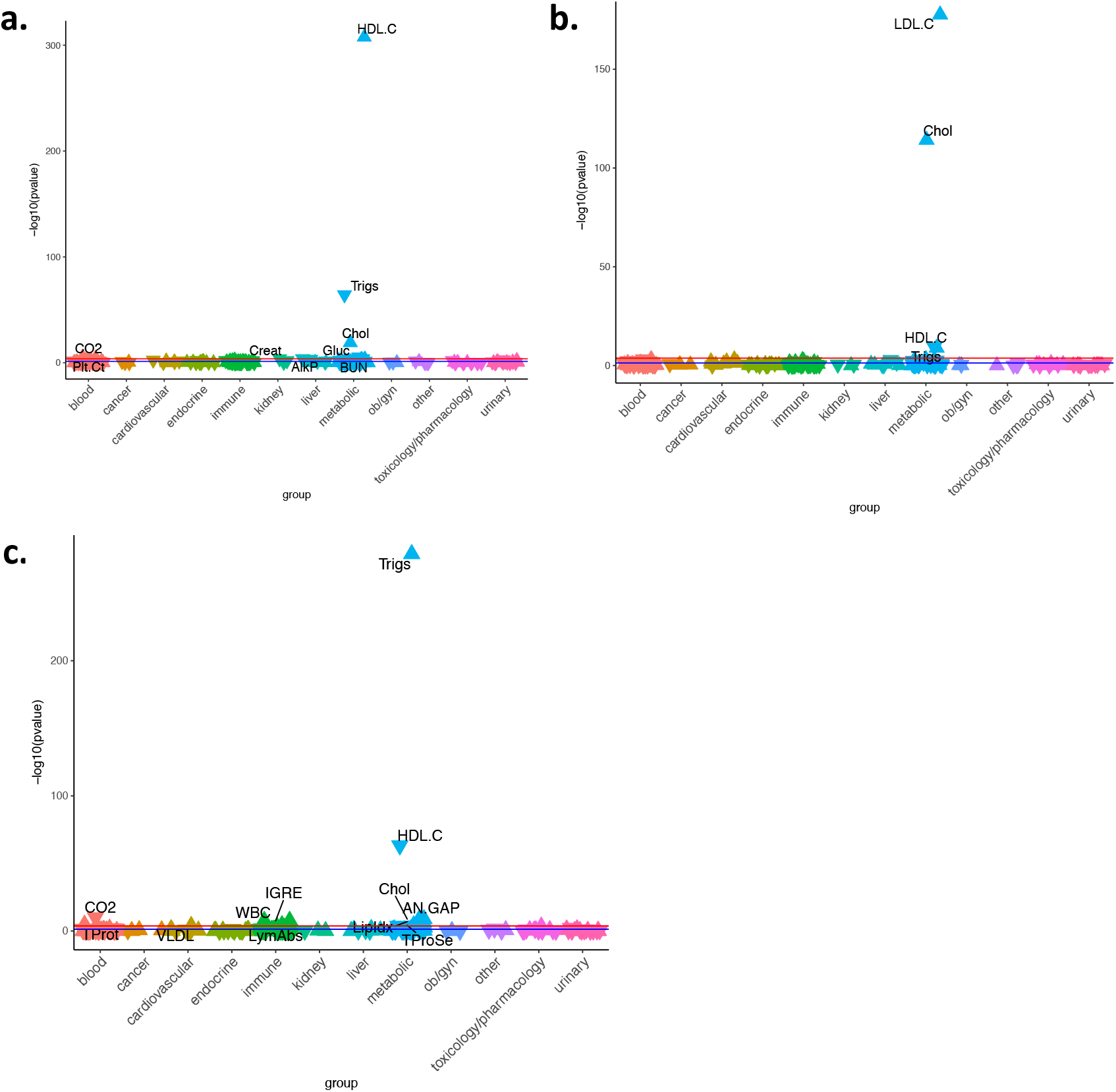
LabWAS of PGS_HDL_ (a), PGS_LDL_ (b), and PGS_TG_ (c). The canonical lab trait is most strongly associated with each PGS. The red line indicates the Bonferroni threshold for statistical significance (p-value of 1.80×10^−4^) and labs with p-values below this threshold labelled. The blue line indicates a p-value of 0.05. Upward triangles indicate that the PGS is associated with increased levels of the lab, while downward triangles indicate an association with reduced levels of the lab.

The LabWAS of LDLPGS showed associations with four lipid labs (Figure 3b, Supplementary Table 7). The most significant association was increased calculated LDL, followed by increased total blood cholesterol, increased TG and decreased HDL.

The LabWAS of TGPGS was associated with five lipids measurements (Figure 3c, Supplementary Table 8), including increased TG, followed by decreased HDL, increased very low-density lipoprotein cholesterol, lipemia index, and total blood cholesterol. Additionally, TGPGS showed associations with three immune labs, two blood protein labs, and two blood composition measurements.

### LabWAS of a Polygenic Score for Coronary Artery Disease

We next sought to recapitulate the risk biomarker profile for CAD through a LabWAS of a CAD_PGS_. The CAD_PGS_ reproduced associations, in the direction of risk, with canonical risk factors for CAD (Figure 4a, Supplementary Table 9), including decreased HDL, increased TG, and increases in two blood glucose measurements, hemoglobin, and glycated hemoglobin (HgbA1C). The CAD_PGS_ also associated with other known biomarkers of cardiovascular health such as increased troponin-I and brain natriuretic peptide (BNP), and lower blood sodium. Finally, the CAD_PGS_ associated with two immune markers and four blood composition measurements.

**Figure 4.**
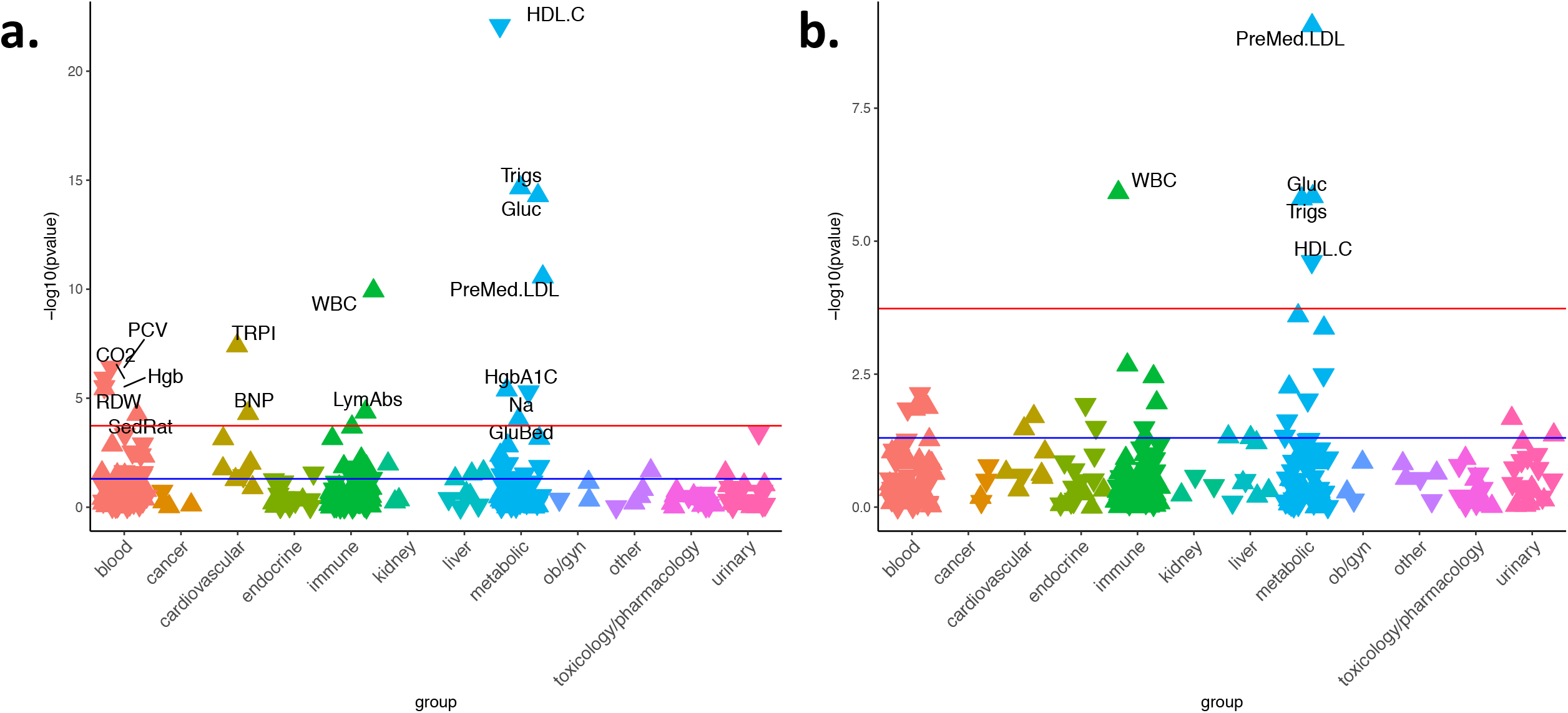
LabWAS of PGS_CAD_. Known lipid biomarkers of CAD are the most strongly associated lab traits (a), even after controlling for a CAD diagnosis (b). The red line indicates the Bonferroni threshold for statistical significance (p-value of 1.80×10^−4^) and labs with p-values below this threshold labelled. The blue line indicates a p-value of 0.05. Upward triangles indicate that the PGS_CAD_ is associated with increased levels of the lab, while downward triangles indicate an association with reduced levels of the lab.

Notably, the CAD_PGS_ was not initially associated with LDL values (p-value=0.78, beta=0.002). The lack of association, however, was attributable to lipid altering medication use and a significant association between the CAD_PGS_ and LDL levels was detected when we restricted to pre-medication values (p = 2.73 × 10^−11^, beta = 0.06).

To determine which biomarkers were explained by the clinical presence of CAD as opposed to just the genetic risk for CAD, we adjusted the LabWAS of CAD_PGS_ for the coronary atherosclerosis phecode (411) (Figure 4b, Supplementary Table 10). Four canonical biomarkers of CAD risk remained associated with CAD_PGS_ including pre-medication LDL, TG, blood glucose, and HDL. Interestingly, the CAD_PGS_ also remained associated with one immune marker (white blood cell count measured in blood).

We also ran a LabWAS of CAD diagnosis (i.e., using CAD cases/control status (Supplementary Material) as the predictor variable after adjusting for sex and median age across the EHR), which revealed the medical comorbidity pattern of CAD. CAD diagnosis was significantly associated with 81 out of 278 labs in our sample (Supplementary Figure 6, Supplementary Table 11), including 24 blood, 19 metabolic, 17 immune, 5 urinary, 6 cardiovascular, 3 liver, 2 endocrine, and 3 kidney measurements).

## Discussion

The results of our study add to a growing body of evidence indicating that lab values from EHRs with linked genetic data can be mined at scale to identify biomarkers for complex disease ^1-5^. Our proof-of-principle analyses focused on lipids and CAD in 66,914 genotyped BioVU patients and revealed that EHR lipid values cleaned using our QualityLab pipeline were genetically comparable to those measured in samples ascertained for research. An exception to this was the weak genetic correlation between LDL measurements in BioVU versus the GLGC, which improved when we considered only pre-medication LDL measurements controlled for CAD or diabetes diagnosis in BioVU. This improvement suggests that information on lipid-lowering medications may be missing in the EHR of some patients, that acute and chronic illness also affect LDL measurements, and the weak genetic correlation could be due to comorbidities present in the BioVU hospital population. In a LabWAS framework, we showed that PGS for lipids associated robustly to the referent lipid, and that the CAD_PGS_ recapitulated associations with known biomarkers, even after adjusting for the disease diagnosis. Future studies could leverage our pipeline not only for studies of existing biomarkers, but also for analysis of rare or understudied complex traits with no known biomarker associations (e.g., psychiatric disorders).

Furthermore, we show that treatments (in this example, lipid-altering medications) can influence the detection of risk biomarkers at the genetic level. Specifically, the CAD_PGS_ was strongly associated with pre-medication median LDL values, but is not associated with combined pre- and post-medication median LDL values. This finding has important and complex implications for the clinical use of polygenic scores. For example, as preventative treatments for complex diseases are adopted (e.g., lipid-altering medications), the risk factors targeted by those treatments (e.g., lipids) are less likely to play a role in the development of disease (e.g., CAD) in future populations. This means that cases ascertained for GWAS of diseases with available preventative treatments, will be enriched for a different set of genetic (and environmental) risk factors because those individuals with risk factors that can be treated may no longer develop the disease. It is therefore important to keep in mind that polygenic scores, while incredibly valuable, are also a snapshot in time of the genetic profile of those with complex disease and thus are highly susceptible to cohort effects in addition to other known sources of technical and experimental artifacts. Additionally, lab associations with CAD case/control status were much broader than the lab associations with CAD_PGS_, revealing the difference between medical comorbidity and genetic pleiotropy.

We present a framework and tools for association studies using clinical lab data in the EHR. The QualityLab pipeline outlines a series of data filters to ensure clean EHR lab data across different patient groups, and while analyses presented herein are filtered for lab values within the range of common population variation, the QualityLab pipeline is easily adaptable. The LabWAS approach then takes these cleaned lab values and tests their association with a single exposure variable (genetic or otherwise). By using polygenic scores as the exposure variable, our LabWAS analyses focused on common population genetic variation, but the method could be used to test any risk factor for its consequences on EHR labs. For example, instead of focusing on central tendencies of the means using population-level data, a user could choose to retain only extreme lab values in pediatric patients to enrich for rare variants of large effect for sequencing studies.

Though the results and approach presented provide an exciting path forward for genetic analysis of EHR-lab data, important limitations should be acknowledged. First, our analyses focused on associations in patients of European ancestry, a necessary constraint of using current GWAS summary statistics, which are almost all from studies of patients of European ancestry. Our BioVU sample, however, included over 10,000 individuals who self-identified as black or African American. As the number of ancestrally diverse GWAS increase, so too will our ability to identify novel biomarkers in different ancestral groups, and the QualityLab pipeline is poised to deliver on these analyses. The QualityLab pipeline could also have more immediate clinical impact on patients of diverse ethnic groups. Ethnicity strongly influences the distribution of lab tests results in healthy people ^31^, but current reference ranges are developed for entire populations, irrespective of ethnic differences. This ignorance of ethnicity could result in under- or over-diagnosis in some patient groups, and developing ethnicity-informed lab reference ranges is low-hanging fruit for precision medicine.

Second, high-throughput analysis of 481 lab traits in our LabWAS required us to prioritize statistical model performance over coefficient interpretability. In our primary analysis, we transformed lab values to fit the normal distribution to improve the performance of the linear regression models^20^. We applied the rank-based inverse normal quantile transformation to all labs, which ensured trait normality by replacing the value of each observation with its quantile from the standard normal distribution. The inverse normal quantile transformation thus preserved the rank ordering of observations, but not the values themselves, and model coefficients therefore are uninterpretable on the original scale. For example, based on our LabWAS results, we are unable to report the change in LDL levels in mg/dL per SD increase in the CAD_PGS_. Multiple testing correction was another statistical challenge inherent to the high-throughput analysis of lab traits. We used the Bonferroni threshold for statistical significance, but this threshold is overly strict because it ignores the correlation between lab tests. Important associations therefore remain to be discovered below the Bonferroni threshold for statistical significance, and identifying correlations between lab traits is a focus of ongoing work.

As EHR resources grow in size, standardized quality control and analysis pipelines will be necessary to compare results across samples. QualityLab and LabWAS provide a starting point for consistent analysis of lab results stored in various EHR systems. A possible limitation of using clinical data from a hospital population is ascertainment of a less healthy population, which could make conclusions less applicable to the general population. However, we demonstrated that EHR-derived lipids are similar to measurements ascertained in traditional cohort studies, providing a basis for more analyses using EHR data ^32^. Overall, QualityLab and LabWAS are scalable programs that can be used to confirm clinical paradigms and discover new relationships between biomarkers and complex traits.

## Data Availability

GWAS data is available for data on the linked Dropbox website.
Code is available here:
QualityLab: https://bitbucket.org/straubp_vandy/quality_labs/
LabWAS: https://bitbucket.org/juliasealock/labwas/

https://www.dropbox.com/sh/w1pbe0jq1bjkpc5/AAAUIdtBgUybE6iHraE8jvp8a?dl=0

## Code Availability

QualityLab: https://bitbucket.org/straubp_vandy/quality_labs/

LabWAS: https://bitbucket.org/juliasealock/labwas/

## Acknowledgements

### Author Contributions

JKD, JMS, GC, and LKD conceived and designed the study. All authors contributed to the acquisition, analysis, or interpretation of data. JKD, JMS, and LKD drafted the manuscript. All authors contributed to revision of the manuscript.

### Funding/Support

JKD is supported by the Canadian Institutes of Health Research (award MFE-142936). JMS is supported by NIH/NIGMS training grant 5T32GM080178-12. JDM is supported by AHA grant 16FTF30130005 and NIH GM130791-01. GC is supported by NIH UL1TR000427 and 1U24CA242637-01. LKD is supported by NIH 1R01MH118233-01, 5U54MD010722-04, 5R01MH113362-03, 1R56MH120736-01.

This project was conducted in part using the resources of the Advanced Computing Center for Research and Education at Vanderbilt University, Nashville, TN. The datasets used for this project were obtained from Vanderbilt University Medical Center’s Synthetic Derivative, which is supported by numerous sources: institutional funding, private agencies, and federal grants. These include the NIH funded Shared Instrumentation Grant S10RR025141; and CTSA grants UL1TR002243, UL1TR000445, and UL1RR024975.

## References

1 Shameer, K. et al. A genome- and phenome-wide association study to identify genetic variants influencing platelet count and volume and their pleiotropic effects. Hum Genet 133, 95–109, doi:10.1007/s00439-013-1355-7 (2014).

2 Hoffmann, T. J. et al. A large electronic-health-record-based genome-wide study of serum lipids. Nat Genet 50, 401–413, doi:10.1038/s41588-018-0064-5 (2018).

3 Verma, A. et al. PheWAS and Beyond: The Landscape of Associations with Medical Diagnoses and Clinical Measures across 38,662 Individuals from Geisinger. Am J Hum Genet 102, 592–608, doi:10.1016/j.ajhg.2018.02.017 (2018).

4 Klarin, D. et al. Genetics of blood lipids among ∼300,000 multi-ethnic participants of the Million Veteran Program. Nat Genet 50, 1514–1523, doi:10.1038/s41588-018-0222-9 (2018).

5 Verma, A. et al. Integrating Clinical Laboratory Measures and Icd-9 Code Diagnoses in Phenome-Wide Association Studies. Pac Symp Biocomput 21, 168–179 (2016).

6 Estiri, H., Klann, J. G. & Murphy, S. N. A clustering approach for detecting implausible observation values in electronic health records data. BMC Med Inform Decis Mak 19, 142, doi:10.1186/s12911-019-0852-6 (2019).

7 Pivovarov, R., Albers, D. J., Sepulveda, J. L. & Elhadad, N. Identifying and mitigating biases in EHR laboratory tests. J Biomed Inform 51, 24–34, doi:10.1016/j.jbi.2014.03.016 (2014).

8 Perrotta, P. L. & Karcher, D. S. Validating Laboratory Results in Electronic Health Records: A College of American Pathologists Q-Probes Study. Arch Pathol Lab Med 140, 926–931, doi:10.5858/arpa.2015-0320-CP (2016).

9 Denny, J. C., Bastarache, L. & Roden, D. M. Phenome-Wide Association Studies as a Tool to Advance Precision Medicine. Annu Rev Genomics Hum Genet 17, 353–373, doi:10.1146/annurev-genom-090314-024956 (2016).

10 Denny, J. C. et al. Systematic comparison of phenome-wide association study of electronic medical record data and genome-wide association study data. Nat Biotechnol 31, 1102–1110, doi:10.1038/nbt.2749 (2013).

11 Pendergrass, S. A. et al. Phenome-wide association study (PheWAS) for detection of pleiotropy within the Population Architecture using Genomics and Epidemiology (PAGE) Network. PLoS Genet 9, e1003087, doi:10.1371/journal.pgen.1003087 (2013).

12 Robinson, J. R., Denny, J. C., Roden, D. M. & Van Driest, S. L. Genome-wide and Phenome-wide Approaches to Understand Variable Drug Actions in Electronic Health Records. Clin Transl Sci 11, 112–122, doi:10.1111/cts.12522 (2018).

13 Roden, D. M. et al. Development of a large-scale de-identified DNA biobank to enable personalized medicine. Clin Pharmacol Ther 84, 362–369, doi:10.1038/clpt.2008.89 (2008).

14 Purcell, S. et al. PLINK: a tool set for whole-genome association and population-based linkage analyses. Am J Hum Genet 81, 559–575, doi:10.1086/519795 (2007).

15 Price, A. L. et al. Principal components analysis corrects for stratification in genome-wide association studies. Nat Genet 38, 904–909, doi:10.1038/ng1847 (2006).

16 Patterson, N., Price, A. L. & Reich, D. Population structure and eigenanalysis. PLoS Genet 2, e190, doi:10.1371/journal.pgen.0020190 (2006).

17 Das, S. et al. Next-generation genotype imputation service and methods. Nat Genet 48, 1284–1287, doi:10.1038/ng.3656 (2016).

18 1000 Genomes Project Consortium et al. A global reference for human genetic variation. Nature 526, 68–74, doi:10.1038/nature15393 (2015).

19 Sofer, T. et al. A fully adjusted two-stage procedure for rank-normalization in genetic association studies. Genet Epidemiol 43, 263–275, doi:10.1002/gepi.22188 (2019).

20 McCaw, Z. R., Lane, J. M., Saxena, R., Redline, S. & Lin, X. Operating characteristics of the rank-based inverse normal transformation for quantitative trait analysis in genome-wide association studies. Biometrics, doi:10.1111/biom.13214 (2019).

21 Yang, J., Lee, S. H., Goddard, M. E. & Visscher, P. M. GCTA: a tool for genome-wide complex trait analysis. Am J Hum Genet 88, 76–82, doi:10.1016/j.ajhg.2010.11.011 (2011).

22 Jiang, L. et al. A resource-efficient tool for mixed model association analysis of large-scale data. Nat Genet 51, 1749–1755, doi:10.1038/s41588-019-0530-8 (2019).

23 Willer, C. J. et al. Discovery and refinement of loci associated with lipid levels. Nat Genet 45, 1274–1283, doi:10.1038/ng.2797 (2013).

24 Bulik-Sullivan, B. K. et al. LD Score regression distinguishes confounding from polygenicity in genome-wide association studies. Nat Genet 47, 291–295, doi:10.1038/ng.3211 (2015).

25 Evans, L. M. et al. Comparison of methods that use whole genome data to estimate the heritability and genetic architecture of complex traits. Nat Genet 50, 737–745, doi:10.1038/s41588-018-0108-x (2018).

26 Bulik-Sullivan, B. et al. An atlas of genetic correlations across human diseases and traits. Nat Genet 47, 1236–1241, doi:10.1038/ng.3406 (2015).

27 Ge, T., Chen, C. Y., Ni, Y., Feng, Y. A. & Smoller, J. W. Polygenic prediction via Bayesian regression and continuous shrinkage priors. Nat Commun 10, 1776, doi:10.1038/s41467-019-09718-5 (2019).

28 Vilhjalmsson, B. J. et al. Modeling Linkage Disequilibrium Increases Accuracy of Polygenic Risk Scores. Am J Hum Genet 97, 576–592, doi:10.1016/j.ajhg.2015.09.001 (2015).

29 Choi, S. W. & O’Reilly, P. F. PRSice-2: Polygenic Risk Score software for biobank-scale data. Gigascience 8, doi:10.1093/gigascience/giz082 (2019).

30 Nikpay, M. et al. A comprehensive 1,000 Genomes-based genome-wide association meta-analysis of coronary artery disease. Nat Genet 47, 1121–1130, doi:10.1038/ng.3396 (2015).

31 Tahmasebi, H., Trajcevski, K., Higgins, V. & Adeli, K. Influence of ethnicity on population reference values for biochemical markers. Crit Rev Clin Lab Sci 55, 359–375, doi:10.1080/10408363.2018.1476455 (2018).

32 Casey, J. A., Schwartz, B. S., Stewart, W. F. & Adler, N. E. Using Electronic Health Records for Population Health Research: A Review of Methods and Applications. Annu Rev Public Health 37, 61–81, doi:10.1146/annurev-publhealth-032315-021353 (2016).

